# Aberrant brain-heart coupling is associated with the severity and prognosis of hypoxic-ischemic brain injury after cardiac arrest

**DOI:** 10.1101/2023.03.13.23287230

**Authors:** Bertrand Hermann, Diego Candia-Rivera, Tarek Sharshar, Martine Gavaret, Jean-Luc Diehl, Alain Cariou, Sarah Benghanem

## Abstract

**Background and Objectives:** Approximately 50% of post-cardiac arrest survivors remain comatose after 72h, a substantial proportion of which will have a poor neurological outcome, predominantly due to irreversible hypoxic-ischemic brain injury. Recent findings in healthy subjects and patients suggested that autonomic nervous system activity measured by brain-heart interactions could be reliable markers of consciousness and cognitive processing. Thus, we hypothesized that brain-heart interactions are associated with the severity of hypoxic-ischemic brain injury and the prognosis of these patients.

**Methods:** In post-cardiac arrest patients still comatose 48h after sedation weaning, brain-heart interaction markers were computed on 5 minutes of continuous EEG/ECG recording using a synthetic data generation model, gathering bidirectional interactions between EEG frequency bands (delta, theta and alpha) and heart-rate variability frequency bands (low and high frequency). The strength and complexity of the interactions were quantified using medians and refined composite multiscale entropy. Primary outcome was the severity of brain injury, assessed by: (i) standardized qualitative EEG classification, (ii) somatosensory evoked potentials (N20), and (iii) neuron-specific enolase levels. Secondary outcome was the 3-month neurological status, assessed by the Cerebral Performance Category score [good (1-2) vs. poor outcome (3-4-5)].

**Results:** Between January 2007 and July 2021, 181 patients [116 males (64%), median age 61 years, age range 49-72 years] were admitted to ICU for a resuscitated cardiac arrest (76% out-of-hospital, 69% non-shockable rhythm). Poor neurological outcome was observed in 134 patients (74%). Qualitative EEG patterns suggesting high severity were associated with a decreased sympatho-vagal balance. Severity of EEG changes were proportional to higher absolute values of brain-to-heart coupling strength (p<2×10^−3^ for all brain-to-heart frequencies) and lower values of complexity (all p-values<0.05 except for alpha-to-low frequency). Brain-to-heart coupling strength was significantly higher in patients with bilateral absent N20 and correlated with neuron-specific enolase levels at day 3. This aberrant brain-to-heart coupling (increased strength, decreased complexity) was also associated with 3-month poor neurological outcome.

**Discussion:** Our results suggest that autonomic dysfunctions may well represent hypoxic-ischemic brain injury post-cardiac arrest pathophysiology. These results open avenues for integrative monitoring of autonomic functioning in critical care patients with potential prognostic applications.

## Introduction

Despite successful resuscitation with the return of spontaneous circulation (ROSC), approximately 50% of patients are still comatose 72h after a cardiac arrest (CA)^1^. A substantial proportion of these comatose patients will present a poor neurological outcome mostly due to irreversible hypoxic-ischemic brain injury (HIBI)^2,3^. The clinical expression of HIBI varies widely, from minimal cognitive impairment to persistent and severe loss of consciousness (i.e., minimally conscious state, vegetative state, or even coma). Those outcomes suggest that the neurological damage causes a disruption of key systems supporting wakefulness and consciousness, such as the ascending reticular system with hypothalamic and thalamic projections to the fronto-parietal cortical network^4^.

Several tools have been developed to assess the extent of brain injury and to guide neuroprognostication^5^. Methods range from the evaluation of brainstem and cortical integration of peripheral sensory information to the assessment of the complex interplay between subcortical and thalamo-cortical loops. The integration of sensory information can be assessed through pupillometry, auditory evoked potentials, or somatosensory evoked potentials (SSEP)^6^. The disrupted interplay between brain structures can be assessed with qualitative^7,8^ and quantitative^9–11^ analysis of EEG activity, and the analysis of axonal and neuronal injury by quantifying serum biomarkers (neuron-specific enolase, NSE), light-chain neurofilament light^12^, and cerebral MRI^13^. However, even with these tools, the prognosis may remain uncertain underlining the need for a multimodal approach and the development of new physiology-grounded prognostic markers.

Recent theoretical developments and experimental results suggested that quantifying the neural monitoring of visceral inputs, through measures of brain-heart interactions, predicts conscious perception^14^. Evidence in patients suffering from disorders of consciousness suggested that measures of brain-heart interactions can detect the presence of consciousness after an acute comatose stage^15–17^. In turn, the brain regulates vital and immune functions through the autonomic nervous system (ANS) comprising both the sympathetic nervous system and parasympathetic nervous system^18^. The ANS plays a key role in homeostasis and allostasis and numerous pieces of evidence suggest a dysfunction of the ANS in critically-ill patients with primary brain involvement such as TBI^19^, temporal and insular epilepsies^20^ or stroke^21^, but also without primary brain lesions^22^ such as in sepsis^23^. In CA patients, a recent study suggested that ANS dysfunction (reflected by impairment of heart-rate variability metrics and decrease of sympathetic tone) was associated with poor outcomes^24^. The neural structures involved in autonomic control include the brainstem, sympathetic and parasympathetic nuclei in the medulla, and several high-order regions, such as the hypothalamus, the amygdala, the insula, and the medial prefrontal cortex. Interestingly, all these brain regions are closely associated with both mechanisms of consciousness and acute responses to stress while being susceptible to HIBI after CA.

We thus hypothesized that altered brain-heart interactions would relate to the severity of HIBI and the outcome of patients resuscitated from CA. In this study, we described brain-heart interactions using a physiological model to describe causality and directionality between ongoing fluctuations in EEG and cardiac sympathetic-vagal oscillations^25,26^ in CA survivors, still comatose at the time of examination (72h after ROSC) according to the severity of HIBI and patients’ neurological outcome at 3 months.

## Material & Methods

### Population

This is a retrospective analysis performed on a prospectively collected database including all consecutive patients admitted in the single tertiary intensive care unit of hospital Cochin in Paris (France) following cardiopulmonary resuscitation and ROSC. For the present analysis, we included adult patients still comatose (as defined by a Glasgow Coma Scale [GCS] ≤ 8 with a GCS motor component < 3 and a Richmond Agitation–Sedation Scale RASS ≤ −4) 48 hours after sedation weaning, who undergone a standard EEG for neuroprognostication. We excluded patients investigated for brain death diagnosis, patients awake before EEG, and patients who died within 48 hours after CA before a reliable neurological examination could be performed.

### Data collection

Data regarding patients’ characteristics, pre-admission and ICU management were collected following the Utstein style. In addition, pre-exposure to beta blockers, ongoing sedation and/or catecholamines at the time of EEG/ECG acquisition were also recorded, as well as length of stay in ICU, vital and neurological status at ICU discharge and cause of death. In a subset of patients, day 3 NSE serum levels and SSEP recordings (at least 48h after sedation discontinuation) were performed and collected.

### Standard Protocol Approvals, Registrations, and Patient Consents

Patients’ next of kin were informed that data were collected for clinical research purposes. Data collection was approved by the Ethics Committee of the French Intensive Care Society (#CESRLF_12-384 and 20–41) and conducted according to French health authorities’ regulations (French Data Protection Authority #MR004_2209691).

### ICU management and neurological prognostication

The management protocol for patients admitted to our ICU after CA has been previously described and did not change throughout the study period^27^. Briefly, in the absence of contraindication, targeted temperature management was immediately started after ICU admission with a target temperature of 32-36°C adapted to hemodynamic tolerance and using an external cooling device for 24 h. A sedation protocol based on the RASS was used according to guidelines, with midazolam and sufentanil before 2014 and the short acting drugs propofol and remifentanil after, titrated to obtain a RASS level of −5 (no response to voice or physical stimulation) before cooling. Sedation was interrupted after rewarming (Supplementary Material A1). Neurological status was evaluated every 3 hours by nurses, and daily by ICU physicians until death or ICU discharge. Awakening was defined as three consecutive RASS scores of at least −2 (patient briefly awakens with eye contact to voice), as previously reported (Ely, JAMA 2003). In patients who were still comatose 72 hours after ROSC and 48 hours after sedation discontinuation, a multimodal prognostication protocol was used, consistent with the 2015 international ERC/ESICM guidelines^28^, including clinical examination, standard EEG, SSEP and NSE at day 3, unchanged during the study period (Supplementary Material A2). Modalities of SSEP recording and interpretation are reported in Supplementary Material A3.

### EEG acquisition and qualitative analysis

Standard 20-minutes EEG (using 13 scalp electrodes: Fp1, Fp2, F7, F8, T3, T4, T5, T6, C3, Cz, C4, O1, O2 in the 10-20 international system, one reference, one ground electrode and two precordial ECG leads) were recorded, with a sample frequency of 250 Hz using Natus Deltamed recording device (Natus, Middleton, USA).

Qualitative analysis of the EEG traces was retrospectively performed by a board-certified electroencephalographer (SB) blinded to the clinical outcome and others prognostic markers. EEGs were analyzed according to the standardized criteria of the American Clinical Neurophysiology Society (ACNS)^29^. Each EEG was classified as highly malignant pattern (suppressed background with or without burst or superimposed periodic pattern), malignant pattern (presence of at least one of the following: abundant generalized periodic or rhythmic discharges, electroencephalographic seizure, discontinuous or low-amplitude background, absence of EEG reactivity) or benign pattern (continuous and reactive pattern, absence of any malignant feature)^7^ reflecting the severity of HIBI.

### EEG and ECG quantitative analysis

ECG were band-pass filtered between 0.5 and 20Hz and EEG were band-pass filtered between 0.5 and 45Hz using Butterworth filters of order 6 and 8 respectively and notch filters were applied at 50Hz and 100Hz. EEG data were re-referenced to an average reference. Independent component analysis (ICA) was applied to EEG (after 1Hz high-pass filtering) using Preconditioned ICA for Real Data (Picard) algorithm and resulting components were visually inspected. ICA components capturing large artifacts (notably eye movements and blinks and cardiac-field artifacts) were subtracted if any and the resulting artifact-free EEG was compared to the original recording to assess the quality of the reconstruction (adequate removal of artifacts and preservation of brain EEG signals). Reconstructed recordings were then visually inspected and periods with remaining artifacts were manually rejected. The first period of 5 minutes of consecutive data without EEG and ECG artifacts was retained in the analysis and further preprocessed as described above.

EEG power spectrum densities were computed using a short-time Fourier transform with a Hamming taper using a sliding window procedure (1 second segments with 75% overlap) to obtain 4 Hz resolution time series of EEG power within the three background EEG frequency bands delta (1-4Hz), theta (4-8Hz), alpha (8-12Hz). Individual EEG channels were denoised using Wavelet thresholding method, with a threshold defined automatically, based on signal’s length and variance^30^.

### Heart-rate variability

Automatic QRS detection was performed using the algorithm described by Elgendi et al^31^. The accuracy of QRS detection was visually inspected and R-peaks were manually corrected if needed to obtain a 5-minutes time series of interbeat intervals (IBI) for each patient. Patients with atrial fibrillation were discarded from the analysis. Given that the computation of the brain-heart interaction relies on the precise timing of both the EEG and IBI time series, no interpolation of ectopic beats was performed. IBI time series intervals were resampled to 4Hz and interpolated using a cubic spline interpolation. Frequency-domain heart-rate variability (HRV) was computed in the low frequency (LF, 0.04-0.15 Hz) and high frequency (HF, 0.15-0.4 Hz). Welch’s method was used to compute the overall power in each frequency band, and the time-varying LF and HF was computed using a smoothed pseudo-Wigner-Ville distribution (a two-dimensional Fourier transform)^32^. Briefly, these two frequency bands have been related respectively to the sympathetic and parasympathetic activities, the LF/HF ratio providing a relative estimation of the sympathovagal balance.

### Heartbeat Evoked Potentials

Heartbeat-evoked potentials (HEPs) aim to capture the transient neural responses triggered by each heartbeat. HEPs were computed by averaging EEG epochs locked to the heartbeat timings, up to 500 ms with respect to the R-peak of the cardiac cycle. Epochs reaching an amplitude > 300 *μ*V were not considered for HEPs computation. Epochs in which the interbeat interval lasted less than 500 ms were not included in the HEPs computation. If more than 20% of EEG epochs were discarded due to the interbeat interval duration, the HEPs computation latency was re-defined to preserve at least 80% of the epochs.

### Brain-heart interplay modeling

Bidirectional brain-heart interactions between EEG within the three frequencies bands of interest and heart rate variability within LF and HF bands were computed using a synthetic data generation (SDG) model^25,33^ (Figure 1). Briefly, the quantification of the functional interplay from the brain to the heart relied on an integral pulse frequency modulation model. The framework aims to model the stimulations to the sinoatrial node that causes the heartbeat generation. Thus, the model considers the interactions between the sympathetic (LF) and parasympathetic (HF) activities, and their respective central control coefficients. The functional interplay from the heart to the brain was estimated through a Markovian synthetic EEG data generation model. The framework models the fluctuations on EEG power on time as an autoregressive process, with an external input (LF or HF).

**Figure 1.**
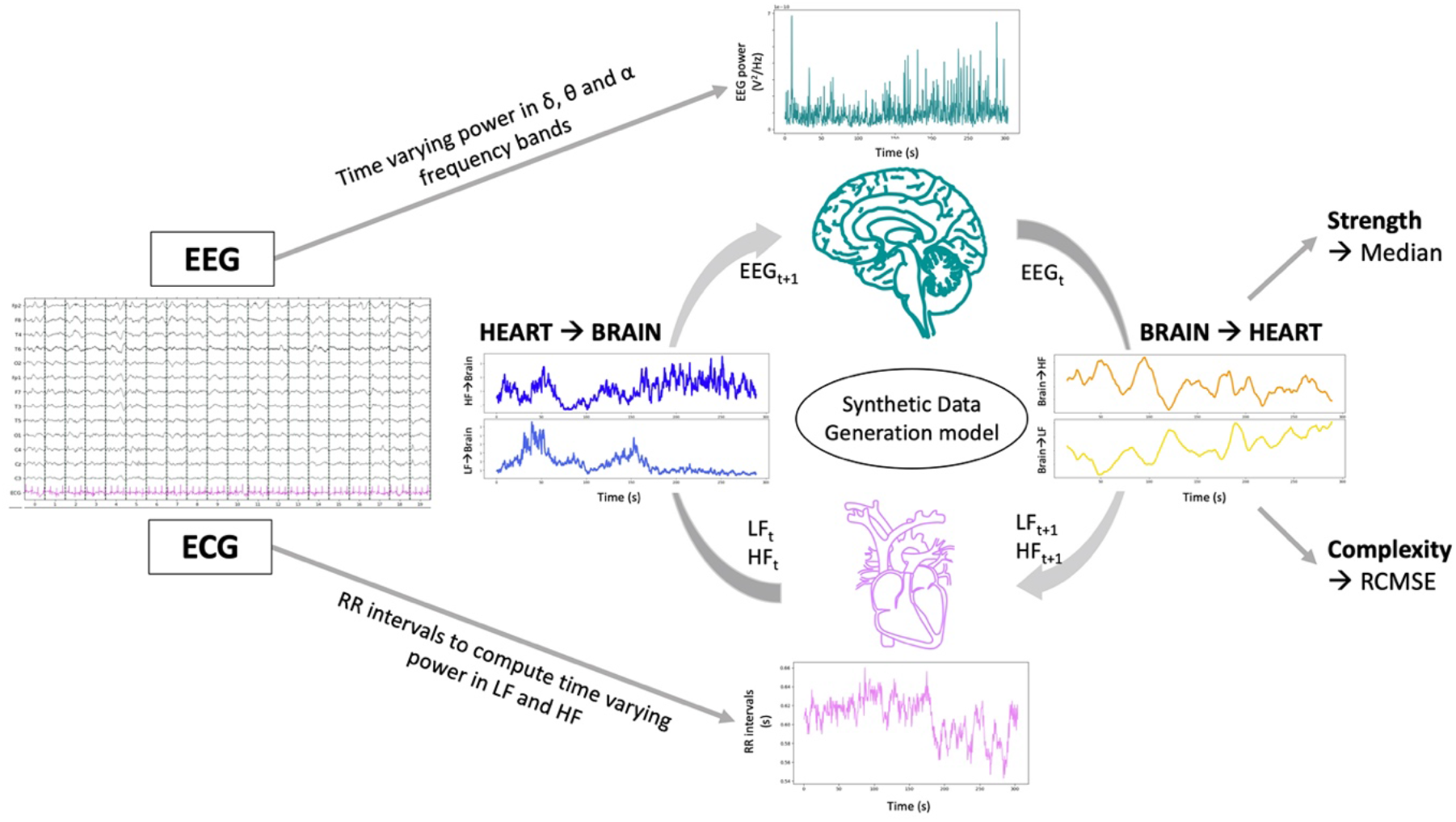
Computation of brain-heart interactions through a synthetic data generation model. Bidirectional brain-heart interactions between the brain within the EEG frequencies of interest (delta, theta, alpha) and the heart within low frequency (LF) and high frequency (HF) bands were computed using a synthetic data generation model. The model estimates the functional interplay between the brain and the heart by assuming a communication loop in which ongoing EEG activity (EEGt) modulates autonomic activity (LFt+1 and HFt+1), while in turn, ongoing autonomic activity (LFt and HFt) modulates EEG activity (EEGt+1). For each EEG-ECG frequency band pair, brain-heart coupling strength and complexity are expressed by the median and refined composite multiscale entropy (RCMSE) of the coupling coefficients over the 5 minutes recording.

The outputs of the model are time-varying coefficients accounting for the brain-heart interplay for each combination of EEG frequency band, LF or HF, and ascending or descending directionality. The computation of brain-heart interplay coefficients was done over EEG and LF/HF power series sampled at 1 Hz, using a 15 s sliding time window. The resulting time-varying coefficients were averaged among channels and on time using the median over the 5 minutes time series to quantify the overall strength of the brain-heart coupling (unitless metrics). Additionally, to better understand high or low coupling coefficients, we also assessed the complexity of the interactions using the refined composite multiscale entropy (RCMSE), an extension of the sample entropy to multiple time scales, with higher values indicating lower predictability and hence higher complexity of the fluctuations of the 5 minutes time series.

### Study endpoints

The primary outcome of the study was the severity of the HIBI as assessed by i) Westhall classification of the EEG according to standardized ACNS terminology (highly malignant, malignant or benign pattern), ii) absence or presence of the N20 SSEP component and iii) serum NSE level at day 3 following ROSC. The other study endpoint was neurological outcome at 3 months, assessed by the ‘best’ Cerebral Performance Category (CPC) level (the most employed scale for assessment of neurological outcome after CA, ranging from 1 to 5). Good neurological outcome was defined as CPC level 1 (no or minimal disability) or 2 (moderate cerebral disability, that is independent for daily living activities), while poor neurological outcome was defined by a CPC of 3 (severe cerebral disability, that is dependent but conscious), 4 (comatose of vegetative state) or 5 (death). We used the ‘best’ CPC observed at 3 months to avoid misclassifying patients reaching CPC 1-2 who subsequently died from non-neurological causes as having a poor neurological outcome due to severe HIBI.

### Statistical analyses

#### Descriptive statistics

Qualitative variables were described using median and interquartile range and compared using Wilcoxon and Kruskal-Wallis tests and categorical variables were described using proportions and compared using Chi-square or Fisher exact tests as appropriate. Correlations between brain-heart interactions markers and NSE levels were performed using the Spearman rho correlation coefficient. Reporting of the study followed the Standards of the STrengthening the Reporting of OBservational studies in Epidemiology (STROBE).

#### Softwares

EEG and ECG data were preprocessed using python and the MNE-python package. Brain-heart coupling was computed using MATLAB R2018b MathWorks) and Fieldtrip Toolbox and statistics were performed using R version 3.6.3 (2020-02-29) (R Core Team (2020). R: A language and environment for statistical computing. R Foundation for Statistical Computing, Vienna, Austria. https://www.R-project.org/) and MATLAB R2018b (MathWorks).

### Data Availability

Clinical and neurophysiological data analyzed in the current study are available from the corresponding author upon reasonable request.

## Results

### Population description

Between January 2007 and July 2021, 240 patients were still comatose at day 3 and underwent EEG for neuroprognostication among whom 181 were included in the analysis (27 patients were discarded due to atrial fibrillation and the remaining patients were not retained in the analysis because of insufficient data quality, see flowchart Figure 2).

**Figure 2.**
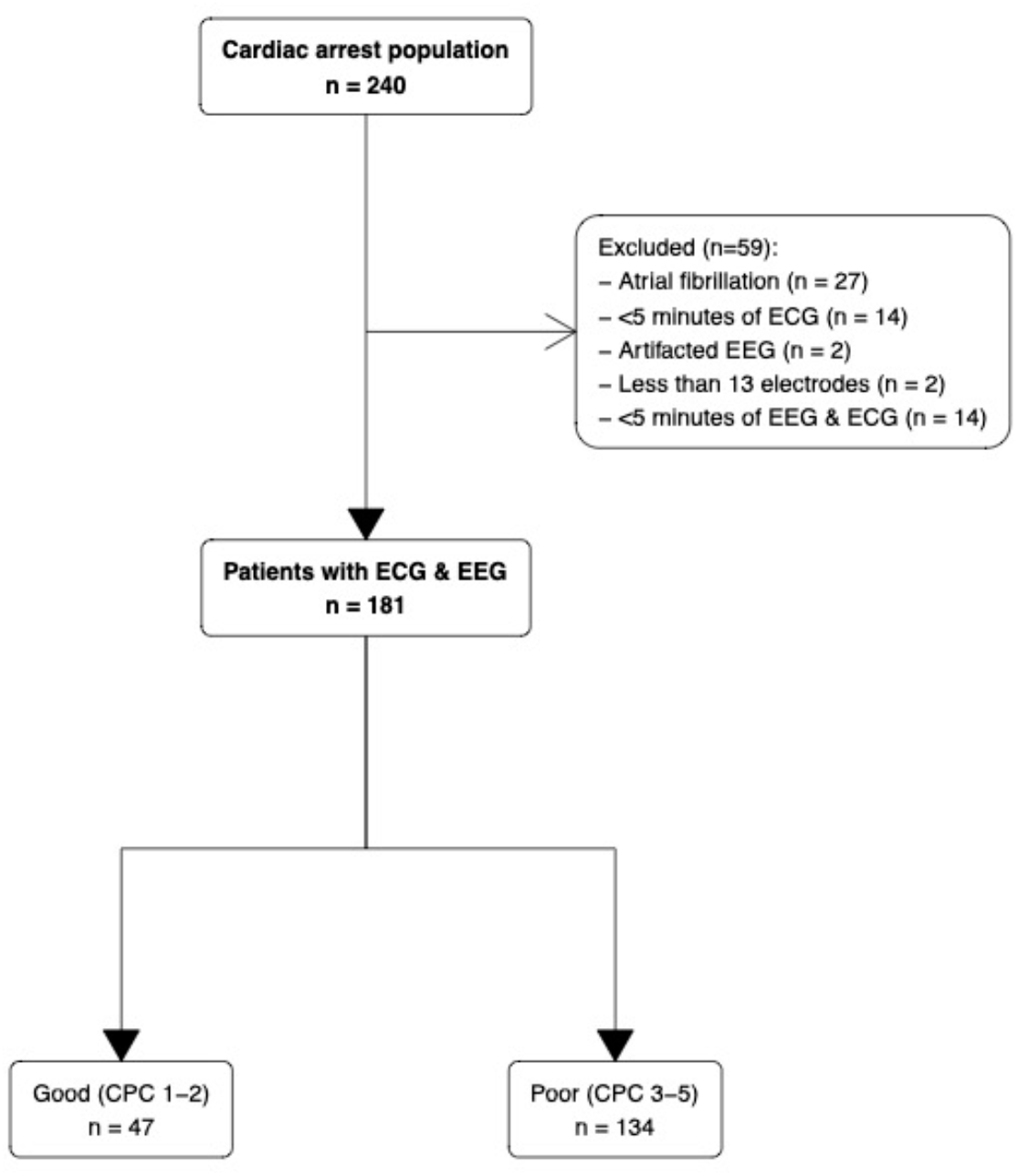
Flowchart.

Patients were mainly male (n=116 (64%)) with a median age of 61 [49-72] years, admitted for out-of-hospital CA (n=138 (69%)) with predominantly non-shockable rhythm. Population characteristics and ICU management are described in Table 1.

**Table 1.** Population characteristics according to the best CPC at 3 months.

| Characteristic | Best CPC at 3 months |  |  | p-value<sup>2</sup> |
| --- | --- | --- | --- | --- |
|  | Overall<br>N = 181<sup>1</sup> | Good<br>N = 47<sup>1</sup> | Poor<br>N = 134<sup>1</sup> |  |
| **Sex (women)** | 65 (36) | 16 (34) | 49 (37) | 0.756 |
| **Age (years)** | 61 [50-72] | 58 [45-66] | 63 [51-73] | 0.077 |
| **Beta blockers before ICU admission (n=178)** | 41 (23) | 8 (17) | 33 (25) | 0.254 |
| **Cardiac arrest location** |  |  |  | 0.740 |
| In-hospital | 43 (24) | 12 (26) | 31 (23) |  |
| Out-of-hospital | 138 (76) | 35 (74) | 103 (77) |  |
| **Witnessed cardiac arrest (n=174)** | 120 (69) | 31 (69) | 89 (69) | 0.990 |
| **Initial rhythm** |  |  |  | 0.063 |
| Asystole | 94 (52) | 18 (38) | 76 (57) |  |
| Ventricular tachycardia/fibrillation | 57 (31) | 22 (47) | 35 (26) |  |
| Pulseless electrical activity | 14 (7.7) | 4 (8.5) | 10 (7.5) |  |
| Unknown | 16 (8.8) | 3 (6.4) | 13 (9.7) |  |
| **No flow (min)** | 3 [0-7] | 1.5 [0-5] | 3.0 [0-7] | 0.213 |
| **Low flow (min) (n=175)** | 18 [10-25] | 15 [7-20] | 20 [12-27] | **0.005** |
| **Hypothermia (n=178)** | 154 (87) | 43 (91) | 111 (85) | 0.245 |
| **ICU mortality** | 129 (71) | 2 (4.3) | 127 (95) | **4e-32** |
| **Delay of EEG (days)** | 3 [2-4] | 2 [1-3] | 3 [2-4] | **0.027** |
| **Sedation during EEG (n=179)** | 48 (27) | 16 (34) | 32 (24) | 0.193 |
| **EEG ACNS standardized classification** |  |  |  | **2e-20** |
| Highly malignant | 41 (23) | 0 (0) | 41 (31) |  |
| Malignant | 92 (51) | 10 (21) | 82 (61) |  |
| Benign | 48 (27) | 37 (79) | 11 (8.2) |  |
| **SSEP N20 presence (n=112)** | 80 (71) | 21 (100) | 59 (65) | **0.001** |
| **NSE day 3 (μmol/L) (n=66)** | 93 [36-259] | 28 [18- 41] | 160 [65-353] | **6e-05** |

<sup>1</sup>n (%); Median [25%-75%]
<sup>2</sup>Pearson's Chi-squared test; Wilcoxon rank sum test; Fisher's exact test

Neurological outcome was unfavorable (CPC 3-4-5) in 134 (74%). EEG recordings were obtained in all patients at a median time of 3 [2-4] days after CA (Table 1).

Standardized qualitative EEG pattern according to the ACNS classification was highly malignant in 41 (23%), malignant in 92 (51%), and benign in 48 (26%). SSEPs were assessed in 112 (62%) patients, among whom 32 (29%) had bilaterally absent N20. A highly malignant pattern was associated with poor outcomes in all cases, as it was for the bilaterally absent N20 pattern. Blood NSE was available in 66 patients, with a median of 28 [18-41] µmol/L and 160 [65-353], p<10^−4^) in good and poor outcome patients, respectively.

### Post CA is characterized by an aberrant brain-to-heart coupling, scaling with HIBI severity

We first investigated the LF/HF ratio according to the severity of HIBI, assessed by the grade of the EEG ACNS classification. LF/HF diminished with EEG patterns of increasing severity, from 1.87 [0.68-3.99] in benign, to 1.24 [0.44-2.67] in malignant and 0.74 [0.26-1.21] in highly malignant EEG patterns (*p*=0.012), suggesting a decreased sympatho-vagal balance in severe HIBI. This seemed to be mainly driven by a decreased LF power although this difference did not reach statistical significance (*p*=0.087).

We then found that both the strength and complexity of the brain-to-heart coupling showed significant associations with the severity of HIBI, assessed by the grade of the EEG ACNS classification. Severity of EEG patterns were proportional to brain-to-heart absolute values of coupling strength (*p*<2×10^−3^ for all couplings between EEG frequencies (i.e., delta, theta or alpha) and ECG frequency activities (i.e., LF and HF), Figure 3A) but inversely proportional to brain-to-heart coupling complexity (all *p*<0.05 except for *alpha-to-LF*, Figure 3B). *Post-hoc* pairwise group comparisons found that the significant differences in coupling strength were mostly differences between patients with highly malignant and either malignant patterns or benign patterns, rather than between malignant and benign patterns. For the differences in coupling complexity, it was the opposite, as they were mainly observed between patients with benign patterns and either malignant or highly malignant patterns (Figure 3 and Supplementary Table 2).

**Figure 3.**
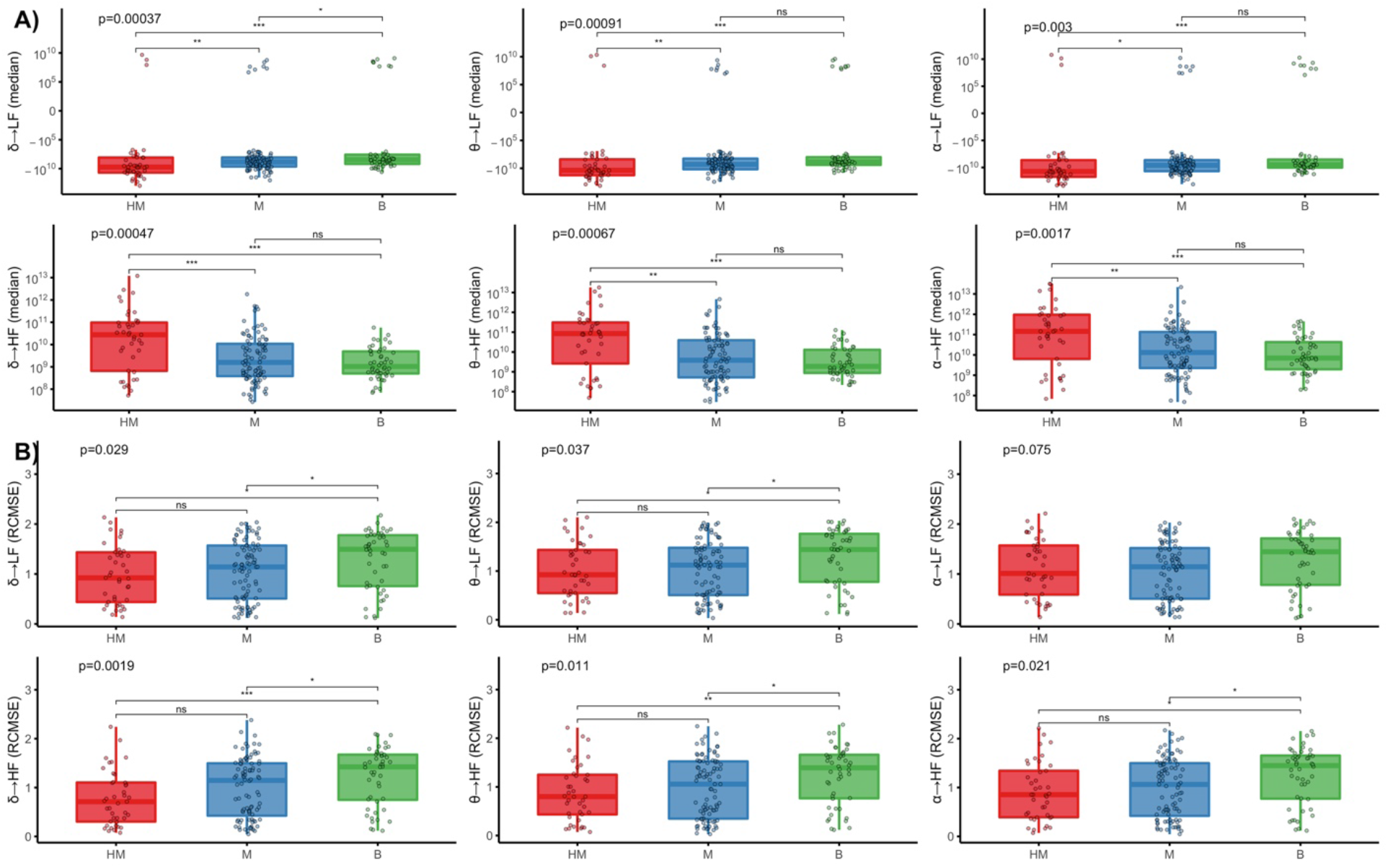
Brain-to-heart coupling strength and complexity according to EEG background ACNS classification. Brain-to-heart coupling strength (median) in **(A)** and complexity (RCMSE) in **(B)** according to EEG background activity following Westhall classification: highly malignant (HM), malignant (M) and benign (B). Overall comparisons were performed using Kruskal-Wallis tests and post-hoc two-by-two group comparisons using Wilcoxon tests. Y-axes for the brain-to-heart coupling strength (median) are in logarithmic scales. *: p≤0.05, **: p≤0.01, ***: p≤0.001, ****: p≤0.0001.

Brain-to-heart coupling strength also scaled with HIBI severity as assessed by two other independent prognostic tools: SSEP and NSE blood levels. First, the median brain-to-heart coupling was higher in patients with bilaterally absent N20 as compared with patients with bilaterally present N20 (Figure 4A and Supplementary Table 3). Second, median brain-to-heart coupling correlated with the NSE blood levels at day 3 after CA, with significant negative correlations between NSE levels and delta-, theta-, and alpha-to-LF (rho between −0.34 and - 0.36 with *p*<0.004) and positive correlations between NSE levels and delta-, theta-, and alpha-to-HF (rho between 0.35 and 0.38 with *p*<0.004, see Figure 4B). No significant differences were observed for brain-to-heart coupling complexity and either SSEP or NSE levels.

**Figure 4.**
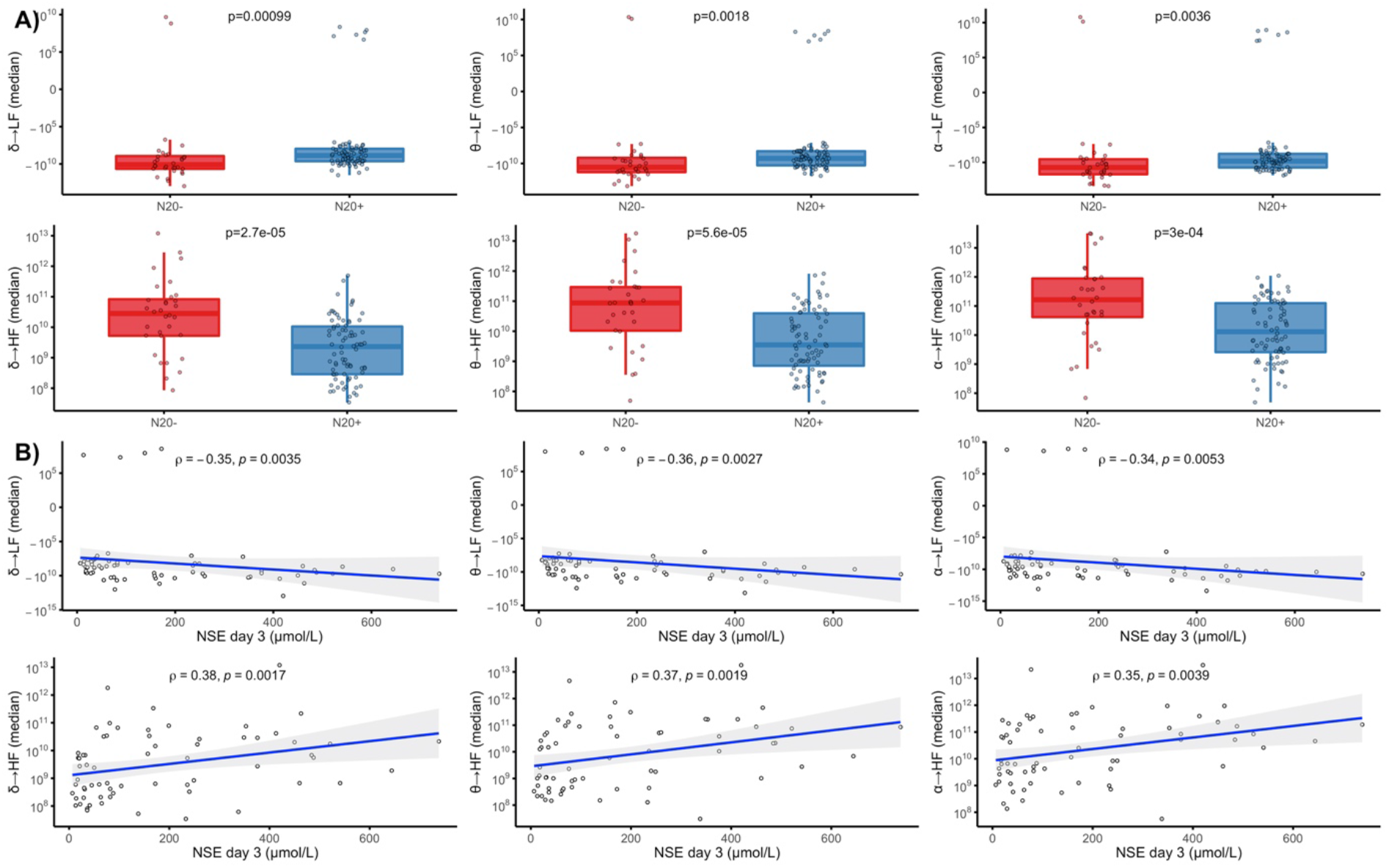
Brain-to-heart coupling strength according to SSEP and day 3 NSE levels. **(A)** Brain-to-heart coupling strength according to the results of the somatosensory evoked potentials (SSEP): bilaterally absent N20 (N20-) versus uni- or bilateral N20 presence (N20+). Groups were compared using Wilcoxon tests. **(B)** Spearman rho (ρ) correlations between median brain-to-heart coupling indices and NSE levels at day 3 after CA. All Y-axes are in logarithmic scales.

### Severe HIBI is also associated with lower strength of the heart-to-brain coupling

To assess bidirectional brain-heart interplay, we also assessed heart-to-brain coupling. While heart-to-brain complexity did not differ according to HIBI severity, heart-to-brain coupling strength was lower in severe HIBI. Indeed, significantly lower values of both LF-to-brain and HF-to-brain were observed with EEG patterns of increasing severity.

*Post-hoc* pairwise comparisons showed significant differences mainly between highly malignant patterns and either malignant or benign patterns (all *p*<0.05, Figure 5A and Supplementary Table 2). Lower values were also observed in patients with bilaterally absent N20 as compared with patients with uni- or bilateral N20 presence (all *p*<0.005, Figure 5B and Supplementary Table 3). Lastly, heart-to-brain coupling also correlated with the NSE levels at day 3 with a lower median coupling coefficient being associated with higher NSE levels (all *p*<0.05 except for the correlation between NSE levels and LF-to alpha, Figure 5C).

**Figure 5.**
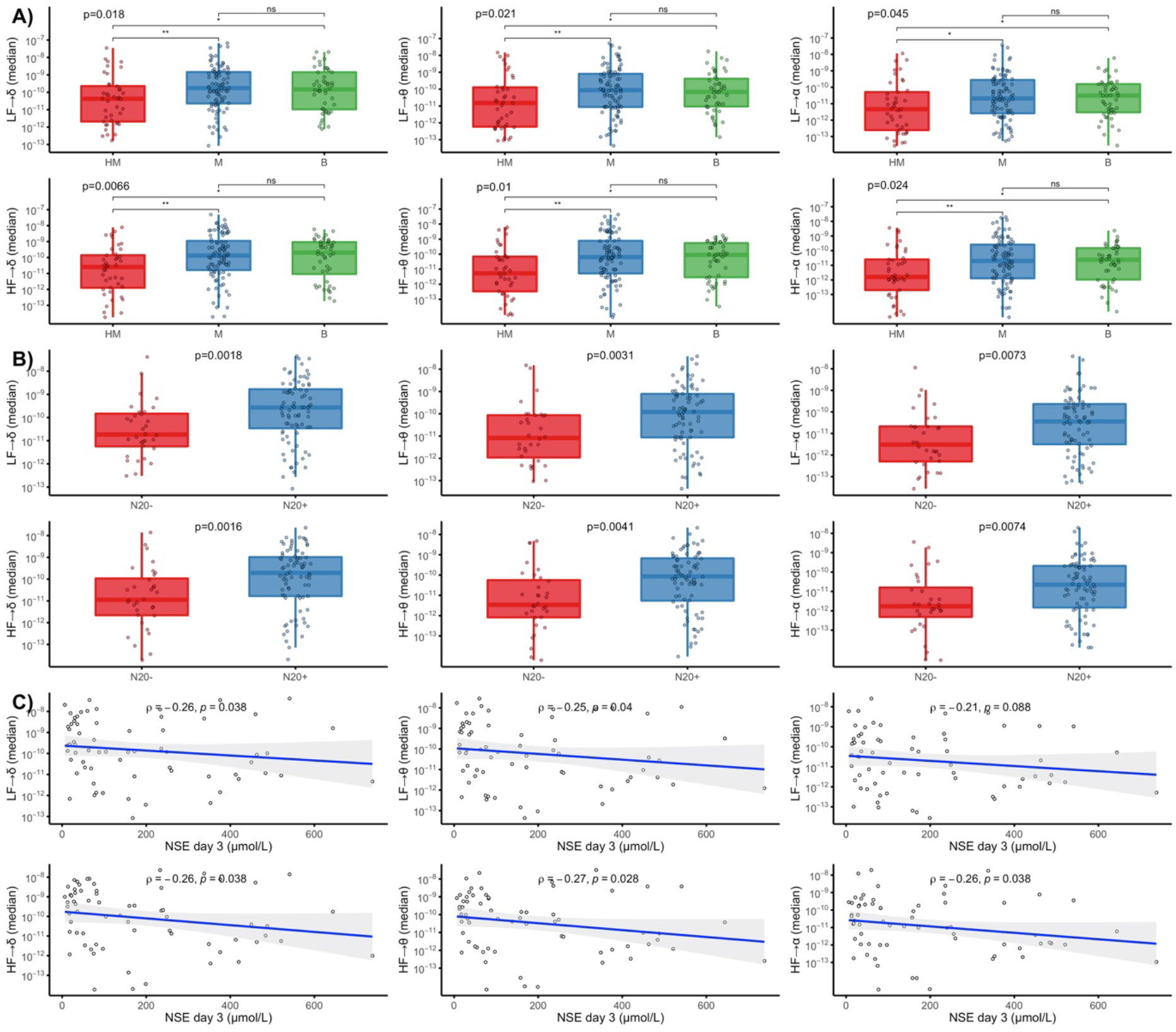
Heart-to-brain coupling strength according to EEG patterns, SSEP, and NSE levels. **(A)** Heart-to-brain coupling strength (median) according to EEG background activity (following Westhall classification: highly malignant (HM), malignant (M) and benign (B)) and, **(B)** according to the results of the somatosensory evoked potentials (SSEP): uni- or bilateral N20 presence (N20+) or bilaterally absent N20 (N20-). **(C)** Spearman rho (ρ) correlations between median heart-to-brain coupling and NSE levels at day 3 after CA. All Y-axes are in logarithmic scales.*: p≤0.05, **: p≤0.01, ***: p≤0.001, ****: p≤0.0001.

### Aberrant brain-to-heart coupling is associated with 3 months neurological outcome

Lastly, we tested if the aberrant brain-heart interactions described above were related to patients’ neurological outcomes.

First, lower LF/HF ratio was observed in patients with poor neurological outcomes as compared to patients with good outcome, 0.97 [0.30-2.44] vs 1.89 [0.76-3.99], *p*=0.017, suggesting a decreased and inversed sympatho-vagal balance in patients with poor neurological outcome. Again, this seemed to be mainly driven by a lower power in the LF band although this difference did not reach statistical significance (0.05 ms^2^ [0.01-0.17] vs 0.08 ms^2^ [0.02-0.41], *p*=0.062) (Supplementary Table 4).

Regarding brain-heart interactions, the strength of brain-to-heart coupling significantly differed between patients with poor and good neurological outcomes especially from delta, theta, and alpha frequency bands to LF (stronger negatives values of median brain-to-heart coupling coefficients in patients with a poor neurological outcome than in patients with a good outcome, all *p*<0.001, Table 2). Brain-to-heart coupling strength also differed between delta-, theta-, and alpha-to-HF (stronger positive median brain-to-heart coupling coefficient in patients with a poor neurological outcome than in patients with a good outcome, all *p*<0.04). As for brain-to-heart coupling complexity, we found that poor outcomes were associated with lower values of RCMSE in both brain-to-LF (delta-to-LF, *p*=0.022, theta-to-LF, *p*=0.016 and alpha-to-LF, *p*=0.036) and brain-to-HF (delta-to-HF *p*=0.015, theta-to-HF, *p*=0.024 and alpha-to-HF, *p*=0.046).

**Table 2.**
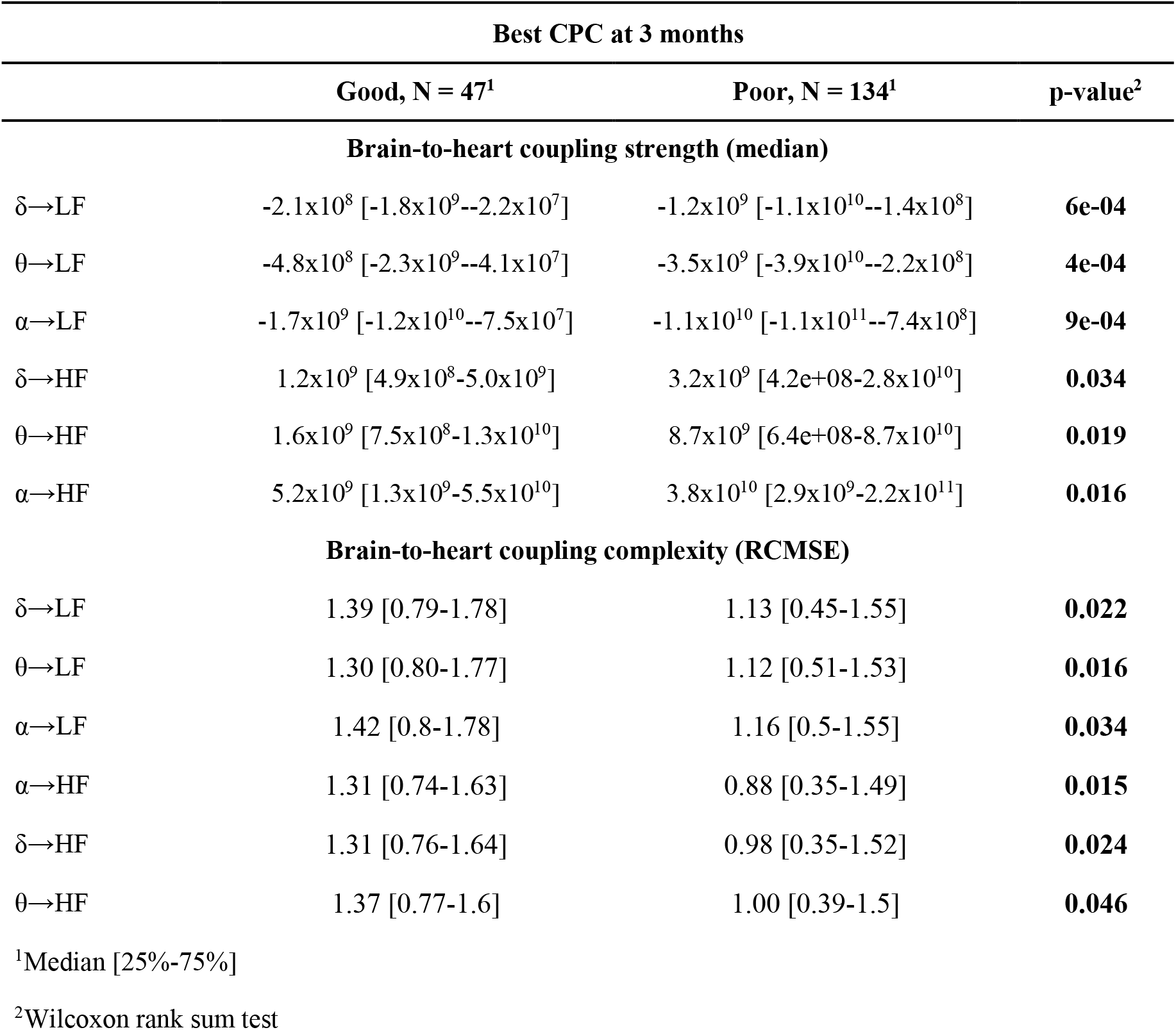
Brain-to-heart coupling strength and complexity according to patients’ outcomes.

Importantly, these differences were probably not explained by the previous exposure to beta-blockers, nor by the infusion of sedation during recording (Supplementary Tables 5 and 6), both being not associated with the neurological outcomes either (*p*=0.25 for beta-blocker exposure and *p*=0.19 for sedation exposure, Table 1). No associations were found between heart-to-brain coupling strength or complexity and neurological outcome. Moreover, the prognostic performances (i.e., specificity, sensitivity, PPV, NPV and AUC) of these markers were poor to moderate, although generally brain-heart coupling markers presented higher AUC than EEG power spectral densities or ECG HRV markers (Supplementary Table 7). Lastly, we did not find difference of heartbeat evoked responses, another frequently used way of assessing ANS function, between patients with poor and good neurological outcomes (Supplementary Figure 2).

## Discussion

Building on existing evidence suggesting that markers on brain-heart interactions can reflect a multisystem dysfunction, we characterized bidirectional brain-heart interactions using a physiologically inspired model in patients resuscitated from cardiac arrest. We found evidence of an autonomic nervous system dysfunction in comatose patients after CA. This disruption of the brain-heart interplay scales with HIBI severity evaluated by EEG patterns, SSEP results, and NSE levels and is characterized by i) a decreased sympatho-vagal balance, ii) an aberrant brain-to-heart coupling (high coupling strength, low coupling complexity), and iii) a low heart-to-brain coupling strength. Lastly, we found that the aberrant brain-to-heart coupling was associated with poor neurological outcomes at 3 months.

ANS dysfunction is a key feature of many critical conditions, with and without direct brainstem lesions. Dysautonomia has thus been described in various etiologies of brain injury such as brain death^34^, stroke^21^, subarachnoid hemorrhage^35^ or TBI^19^ with symptoms ranging from the disappearance of the vasomotor tone, cardiac arrhythmias, paroxysmal sympathetic hyperactivity^36^ sometimes leading to an impairment of myocardial contractility. Interestingly this dysfunction is also described in non-brain-injured critically ill patients and is associated with ICU mortality^23^. So far, the assessment of ANS dysfunction mostly relied on various HRV metrics notably in the frequency domain with two main frequency bands of interest in short recordings: HF band (i.e., 0.15 to 0.4 Hz) predominantly reflecting the parasympathetic tone, and LF (i.e., 0.04 to 0.15 Hz) primarily mediated by sympathetic activity, although both metrics are susceptible to changes in parasympathetic and sympathetic activity^37^. However, so far, studies have either focused on HRV metrics alone^24^ or the cortical integration of heart signals using HEPs^38,39^. We here show that the study of bilateral brain-heart interactions is more comprehensive in evaluating ANS function after CA by providing mechanistic insights into the pathophysiology of HIBI. It also seems to outperform the traditional HEP methods, as HEPs did not differ according to patients’ outcomes in our study. Indeed, using an innovative modeling approach for the assessment of bidirectional brain-heart interplay, to our best knowledge, we show for the first time that HIBI also induces an ANS dysfunction, which is both related to the severity of the injury and the 3-month neurological outcome of patients.

This ANS dysfunction is characterized by a decreased and sometimes inversed sympatho-vagal balance in severe HIBI, suggesting a loss of sympathetic tone. This loss of sympathetic tone was previously described in CA survivors^24^ and has been linked to loss of consciousness in patients with various neurological disorders^40^. Here we show that this reduced sympathetic tone and/or sympatho-vagal balance could be related to a pathologically increased coupling strength from the brain to the heart, with notably stronger negative coupling between EEG power and LF, and a stronger positive coupling between EEG power and HF. This overall increase in coupling strength is also associated with reduced complexity of the brain-to-heart interaction. Taken together, this aberrant coupling is suggestive of a pathological excessive and disproportionate brain-to-heart coupling as observed in loss of consciousness following epilepsy^41^ and general anesthesia^42,43^. This aberrant coupling could reflect the overall slowing, reduction of power and loss of complexity of brain and heart activities. This disproportionate activity has also been observed in rats, during CA^44^ and also before CA, in a recent model of hypoxic CA. In this model, a surge of cortico-cardiac functional and effective connectivity followed hypoxia and preceded the onset of ventricular fibrillation^44^. Another potential relationship in the correlation between brain-heart aberrant coupling and poor outcome is the generalized reduction of physiological activity in both the central and autonomic nervous systems, which could be non-functional. Different animal models have shown that towards the near-death, baseline (intracranial) EEG and heart rate are reduced in parallel^65,66^. These effects are reported in human near-death case reports, where the brain’s cessation of activity occurs in parallel or after the autonomic cessation^39,45^.

On the other hand, this disruption of brain-heart interplay was characterized by a lower heart-to-brain coupling strength and complexity in patients with severe HIBI, reflecting the impairment of bottom-up signaling and integration of peripheral signal. The integration of visceral signals seems to be a hallmark in conscious processing, as evidenced by a recently growing body of literature on perceptual awareness tasks in healthy subjects^14,46^. The potential role of visceral signals in consciousness was tested and confirmed in patients suffering from disorders of consciousness^15,16^.

We thus show that both directions of the brain-heart interplay are impaired in CA survivors with a dose-effect relationship with the severity of HIBI, as assessed by EEG background patterns, results of N20 on SSEP, and serum NSE levels. This multimodal approach to patients’ prognosis with concordant findings from different complementary prognostic markers^47^ and the dose-effect relationship considerably strengthens the validity of our findings and suggests that the degree of ANS dysfunction is probably related to the burden of anoxo-ischemic lesions^48^ which are not uniformly distributed within the brain^1,49^. Unfortunately, brain-imaging or pupillometry data were not available in this cohort to precisely document the regions involved in this aberrant coupling. Notably, it is unknown if this aberrant coupling is due to direct lesions to the brainstem autonomic centers or high-order cortical regions involved in autonomic control. Indeed, in a large post-mortem study, Endisch *et al*. described a predominance of neuronal death in neocortical areas and hippocampus, while the brainstem seemed to be less sensitive to anoxo-ischemic injuries^48^, a finding consistent with the usual preservation of brainstem auditory evoked potentials after CA, even in case of severe HIBI^50^.

Although scaling with HIBI severity and associated with 3 months prognosis, the prognostic performances of the brain-heart and HRV metrics were poor to moderate in comparison to prognostic markers used in clinical practices (i.e., EEG, SSEP and NSE) or to recently published data of early HRV metrics in CA patient during hypothermia^24^. First, our recordings are not as early (3 days in median vs. in the first 24h), nor as long (which could lead to a less robust estimation of LF). We also did not assess other frequencies or non-linear domain HRV metrics as we specifically chose to focus on the brain-heart interplay. It should also be noted that our study is a retrospective analysis of a prospectively collected database. Although our population is homogeneous and treated in the same way throughout the entire period this could have biased the follow-up of patients. ANS could also be influenced or modified by sedation or exposure to beta-blockers, yet it is important to highlight that our results suggest that brain-heart interplay does not differ depending on exposure to these different potential confounders. Lastly, although it is difficult to demonstrate and it was not the main aim of our study, it is still possible that this ANS dysfunction predisposes to several short or long-term complications such as cardiac arrhythmia, hemodynamic failure, maladaptive immune response, and cognitive sequelae in survivors.

## Conclusion

In this study, we investigated the autonomic nervous system function following cardiac arrest through an innovative modeling approach of the bidirectional brain-heart interplay. We showed that post-cardiac arrest survivors exhibit an aberrant brain-heart coupling characterized by an excessive and disproportionate brain-to-heart coupling (increased strength, decreased complexity) and a decreased heart-to-brain coupling, scaling with the severity of HIBI. This aberrant coupling is also associated with patients’ neurological outcome at 3 months. Our results contribute to a better understanding of HIBI pathophysiology and open avenue for an integrative monitoring of the autonomic system functioning in critically patients with potential prognostic applications.

