## Supplementary Material for "Aberrant brain-heart coupling is associated with the severity and prognosis of hypoxic-ischemic brain injury after cardiac arrest"

A. **SUPPLEMENTARY METHODS**
   1. **ICU management protocol after cardiac arrest**
   2. **Neurological prognostication algorithm and criteria for withdrawal of life sustaining therapy**
   3. **Somatosensory evoked potential recordings and interpretation**
B. **SUPPLEMENTARY RESULTS**
   1. **ECG heart-rate variability, EEG power spectral densities and brain-heart coupling strength and complexity according to EEG ACNS classification and SSEP results (Supplementary Tables 2 and 3)**
   2. **ECG heart-rate variability and EEG power spectral densities according to neurological outcome (Supplementary Table 4)**
   3. **Brain-to-heart strength and complexity according to previous beta-blocker exposure and to sedation during EEG recording (Supplementary Tables 5 and 6)**
   4. **Prognostic performances of ECG heart-rate variability, EEG power spectral densities and brain-heart interactions markers (Supplementary Table 7)**
   5. **Heartbeat Evoked Potentials according to outcome (Supplementary Figure 2)**
A. **SUPPLEMENTARY METHODS**
1. **ICU management protocol after cardiac arrest**

Immediately after OHCA and when no obviously other etiology than cardiac origin was retained, all patients had coronary angiography followed by percutaneous coronary intervention if needed. When coronary angiography was inconclusive, brain and angio-thoracic CT scan were performed to identify a reversible cause of arrest. TTM was initiated immediately at ICU admission using external cooling by forced cold air during the first 24 hours to obtain a target temperature between 32 and 34°C as recommended and subsequently rewarmed to 36°C at a rate of 0.3°C/h. Renal replacement therapy was initiated in case of severe metabolic acidosis and/or in case of life-threatening hyperkalemia, defined as a blood potassium level higher than 6 mmol.L^-1^. A mean arterial blood pressure (MAP) between 65 and 75 mmHg was targeted during the ICU stay. Post-resuscitation shock was defined as a MAP<60 mmHg or a systolic blood pressure<90 mmHg sustained for more than six hours after ROSC, despite adequate fluid loading, and requiring norepinephrine or epinephrine infusion. Clinical and non-convulsive seizures were treated with antiepileptic drugs (phenytoin, phosphenytoine, levetiracetam, valproate or phenobarbital). During the first 48 hours of ICU stay, treatments were adapted to maintain homeostasis with glucose control, normocapnia, inspired fraction of O_2_ titrated for arterial saturation of 94-98%, mean arterial pressure target to 65-70mmHg, and hemoglobin level over 7 g.dL^-1^.

Concerning sedation regimen, before 2014 we used midazolam and fentanyl with dose titrated to RASS -5 (no response to voice or physical stimulation). For midazolam the infusion starts at 4 mg.h^-1^ and the rate is increased by 1mg.h^-1^ if RASS -4 or by 2mg.h^-1^ if RASS is -3 or more after a bolus of 2 mg. Fentanyl is started at 0,7 μg.kg^-1^.h^-1^ and the titration is made with steps of 25 μg.h^-1^. When the goal is reached we systematically use vecuronium or atracurium to neuromuscular blocking according to train of four responses with 1 or 2 responses. Sedation was interrupted after rewarming and a train of four responses with 4 responses, ensuring that RASS was assessed only after clearance of NMB. In 2014, considering the high incidence of delayed awakening with midazolam-fentanyl, we have decided to change our sedation regimen towards short-acting-drugs. We used after 2014 a sedative protocol with propofol and remifentanil with dose titrated to RASS -5 (no response to voice or physical stimulation). For propofol the infusion starts at 1 mg.kg^-1^.h^-1^ and the rate is increased by 0.1 mg.kg^-1^.h^-1^ with a maximal dose of 4 mg.kg^-1^.h-^1^. Remifentanil is started at 10 μg.kg^-1^.h^-1^ and the titration is made with steps of 1 μg.kg^-1^.h^-1^ with a maximal dose of 18 μg.kg^-1^.h^-1^. When the sedation goal is reached shivering are appraised according to bedside shivering assessment scale (BSAS) every 3 hours, with a goal of 0 (no shivering). If goal is not achieved a bolus of Atracurium 0.4 mg.kg^-1^ is given, after two bolus in two hours, an infusion of 0.3 mg.kg^-1^.h^-1^ is started. The titration is made with steps of 0.15 mg.kg^-1^.h^-1^. Sedation was interrupted after rewarming and a train of four responses with 4 responses, ensuring that RASS was assessed only after clearance of NMB.

2. **Neurological prognostication algorithm and criteria for withdrawal of life sustaining therapy**

After the initial period of TTM and rewarming, neurological outcome is assessed daily in every patient by ICU physicians until death or ICU discharge. At 48 hours after discontinuation of sedation, in patients who do not awake, GCS, pupillary and corneal reflexes are assessed and an SSEP/EEG are performed. Due to inclusion period (2019-2021), we based our neuroprognostication algorithm on post resuscitation care guidelines published on 2015 (see below). When two or more of the following conditions were present: 1) bilaterally absent pupillary and corneal reflexes; 2) bilaterally absent N20 SSEP responses or 3) a refractory status epilepticus, suppression or burst suppression on EEG recording, and no clinical evidence suggested prolonged sedation, an ethic meeting between all team members (intensivists, nurses, neurologist, therapist), was hold to possibly decide WLST. Conversely, when major predictors of poor outcome were not present (i.e., patients with N20 potentials and cranial reflexes preserved, motor GCS more than 2), decisions to withhold or withdraw life-support therapies were systematically delayed in order to search for a confounding factor (sepsis, remaining sedative drug effect, inter-current disease process, other neurological disease). After this additional delay, an ethic meeting is held to incorporate all prognostic variables in the decision. This decision could be either to withhold or withdraw life-support therapies.

WLST was always decided after a collegial meeting. All deaths associated with end-of-life decisions occurred during the ICU stay.

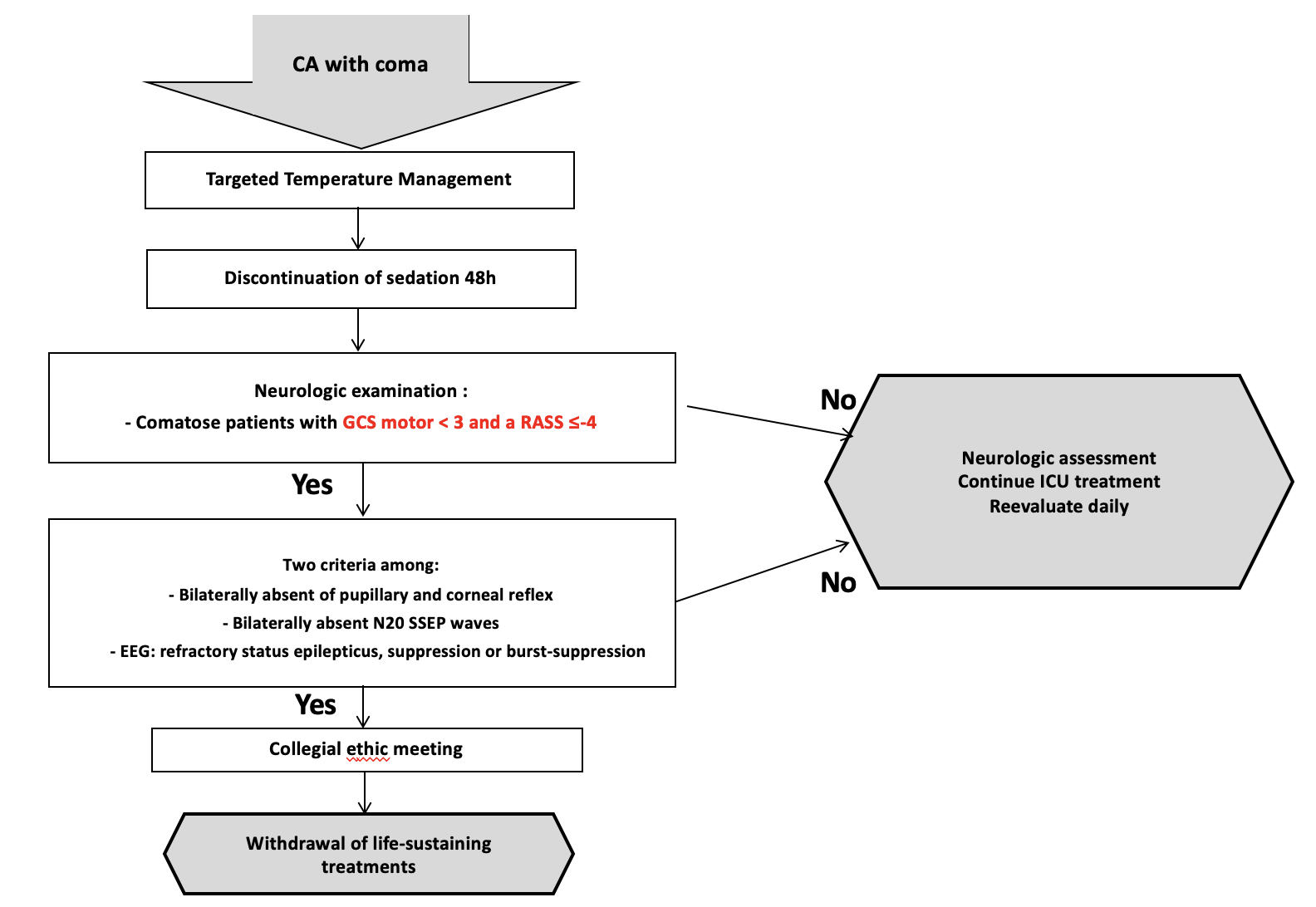

***Supplementary Figure 1.*** ***Decision algorithm for post-resuscitation patients Adapted from Nolan et al, Resuscitation and ICM 2015[14]***

3. **Somatosensory evoked potential recordings and interpretation**

SSEP recordings were made using Deltamed Coherence (Natus, Middleton, USA). SSEP were recorded in patients still comatose 72 hours after ROSC and 48 hours after sedation discontinuation. The SSEP was measured after stimulation of the right and left median nerve using a bipolar surface electrode at the wrist. Stimulation intensity was adjusted to produce visible thumb twitches; if neuromuscular blocking agents were administered, Erb amplitudes were used instead. Monophasic rectangular-wave 200 ms stimulus pulses were delivered. Stimulus frequency was set at 2-3 Hz. Poststimulus recording lasted 50 ms (bandwidth: 30Hz _ 3 kHz, sampling frequency: 50 kHz). Two or three sets of >300–1000 responses were averaged. Surface electrodes were positioned at Erb’s points. Needle electrodes were used for scalp derivations: 2 cm posterior to C3 and C4 (C3’ and C4’). N9 (peripheral), N13 and N20 (cortical) responses were recorded. N9 peripheral responses corresponded to Erb's point ipsilateral to the stimulation versus reference electrode at contralateral Erb's point. For cortical responses, a bi-parietal montage (active electrode contralateral to the stimulation versus reference electrode ipsilateral to the stimulation) was used.

N20 were deemed interpretable if at least 2 peripheral (N9), 2 spinal peak (N13) and cortical recordings (N20-P25) per side were bilaterally reproducible and if a noise level below 0.25 mV in all 4 cortical recordings had been achieved. Noise level was determined 5–10 ms after stimulation to exclude stimulation artifacts. The noise level was determined automatically and visually checked. Digitalized SSEPs were reevaluated blinded to patients’ outcome by an expert electrophysiologist.

B. **SUPPLEMENTARY RESULTS**
1. ***ECG heart-rate variability, EEG power spectral densities and brain-heart coupling strength and complexity according to EEG ACNS classification and SSEP results (Supplementary Tables 2 and 3)***

|  | **EEG ACNS classification** | | |  |
| --- | --- | --- | --- | --- |
| **Characteristic** | **Highly malignant,  N = 41^1^** | **Malignant,  N = 92^1^** | **Benign,  N = 48^1^** | **p-value^2^** |
| **ECG Heart Rate Variability** | | | | |
| **LF (ms²/Hz)** | 0.03 [0.00-0.13] | 0.05 [0.01-0.18] | 0.06 [0.02-0.30] | 0.087 |
| **HF (ms²/Hz)** | 0.03 [0.01-0.11] | 0.03 [0.01-0.16] | 0.04 [0.01-0.15] | 0.562 |
| **LF/HF** | 0.74 [0.26-1.21] | 1.24 [0.44-2.67] | 1.87 [0.68-3.99] | **0.012** |
| **EEG Power Spectral Density** | | | | |
| **δ (V²/Hz)** | 3.6x10^-12^ [1.3x10^-12^-9.2x10^-11^] | 8.7x10^-11^ [1.1x10^-11^-3.0x10^-10^] | 7.0x10^-11^ [3.5x10^-11^-1.5x10^10^] | **1e-04** |
| **θ (V²/Hz)** | 1.5x10^-12^ [4.0x10-13-5.0x10-11] | 3.6x10^-11^ [3.1x10^-12^-1.7x10^-10^] | 3.4x10^-11^ [2.0x10^-11^-5.9x10^-11^] | **2e-04** |
| **α (V²/Hz)** | 9.8x10^-13^ [1.4x10^-13^-2.1x10^-11^] | 8.6x10^-12^ [1.1x10^-12^-3.7x10^-11^] | 1.1x10^-11^ [5.5x10^-12^-2.6x10^-11^] | **3e-04** |
| **Brain-to-heart coupling strength (median)** | | | | |
| **δ→LF** | -5.5x10^9^ [-4.8x10^10^--1.2x10^8^] | -7.0x10^8^ [-4.4x10^9^--1.2x10^8^] | -2.8x10^8^ [-1.5x10^9^--3.8x10^7^] | **4e-04** |
| **θ→LF** | -2.3x10^10^ [-1.8x10^11^--2.5x10^8^] | -2.0x10^9^ [-1.5x10^10^--1.8x10^8^] | -7.2x10^8^ [-2.4x10^9^--1.0x10^8^] | **9e-04** |
| **α→LF** | -5.4x10^10^ [-5.2x1011--4.5x10^8^] | -4.0x10^9^ [-4.8x10^10^--4.4x10^8^] | -3.1x10^9^ [-1.1x10^10^--3.8x10^8^] | **0.003** |
| **δ→HF** | 2.8x10^10^ [6.7x10^8^-9.7x10^10^] | 1.6x10^9^ [3.9x10^8^-1.1x10^10^] | 1.1x10^9^ [5.1x108-4.9x10^9^] | **5e-04** |
| **θ→HF** | 8.5x10^10^ [2.6x10^9^-3.1x10^11^] | 3.9x10^9^ [5.3x10^8^-3.9x10^10^] | 1.9x10^9^ [8.8x10^8^-1.3x10^10^] | **7e-04** |
| **α→HF** | 1.4x10^11^  [6.3x10^9^-9.5x10^11^] | 1.3x10^10^ [2.3x10^9^-1.3x10^11^] | 7.0x10^9^ [2.0x10^9^-4.2x10^10^] | **0.002** |
| **Brain-to-heart coupling complexity (RCMSE)** | | | | |
| **δ→LF** | 0.92 [0.44-1.43] | 1.14 [0.51-1.57] | 1.49 [0.76-1.78] | **0.029** |
| **θ→LF** | 0.92 [0.55-1.43] | 1.12 [0.51-1.48] | 1.44 [0.78-1.76] | **0.037** |
| **α→LF** | 1.01 [0.59-1.57] | 1.14 [0.51-1.52] | 1.45 [0.78-1.71] | 0.075 |
| **δ→HF** | 0.71 [0.30-1.10] | 1.15 [0.43-1.50] | 1.42 [0.75-1.67] | **0.002** |
| **θ→HF** | 0.80 [0.44-1.25] | 1.06 [0.35-1.52] | 1.39 [0.77-1.66] | **0.011** |
| **α→HF** | 0.85 [0.39-1.34] | 1.06 [0.42-1.50] | 1.45 [0.77-1.65] | **0.021** |
| **Heart-to-brain coupling strength (median)** | | | | |
| **LF→δ** | 4.2x10^-11^  [2.1x10-12-2.2x10^-10^] | 1.7x10^-10^  [2.3x10^-11^-1.4x10^-9^] | 1.5x10^-10^ [1.1x10^-11^-1.4x10^-9^] | **0.018** |
| **LF→θ** | 1.5x10^-11^ [5.9x10^-13^-1.3x10^-10^] | 8.7x10^-11^ [8.8x10^-12^-8.0x10^-10^] | 6.8x10^-11^ [9.5x10^-12^-4.2x10^-10^] | **0.021** |
| **LF→α** | 4.6x10^-12^ [2.5x10^-13^-5.1x10^-11^] | 2.1x10^-11^ [2.6x10^-12^-2.8x10^-10^] | 3.1x10^-11^ [3.0x10^-12^-1.5x10^-10^] | **0.045** |
| **HF→δ** | 2.6x10^-11^ [1.3x10^-12^-1.4x10^-10^] | 1.3x10^-10^ [1.7x10^-11^-1.1x10^-9^] | 2.1x10^-10^ [1.1x10^-11^1-9.2x10^-10^] | **0.007** |
| **HF→θ** | 5.3x10^-12^ [3.4x10^-13^-6.9x10^-11^] | 6.4x10^-11^ [5.3x10^-12^-7.6x10^-10^] | 8.9x10^-11^ [2.9x10^-12^-5.4x10^-10^] | **0.010** |
| **HF→α** | 1.6x10^-12^ [2.0x10^-13^-2.5x10^-11^] | 2.0x10^-11^ [1.3x10^-12^-2.6x10^-10^] | 2.4x10^-11^ [1.1x10^-12^-1.5x10^-10^] | **0.024** |
| **Heart-to-brain coupling complexity (RCMSE)** | | | | |
| **LF→δ** | 1.35 [1.04-1.74] | 1.42 [1.17-1.64] | 1.33 [1.17-1.57] | 0.658 |
| **LF→θ** | 1.38 [1.07-1.71] | 1.44 [1.14-1.63] | 1.41 [1.16-1.64] | 0.956 |
| **LF→α** | 1.38 [1.06-1.69] | 1.43 [1.06-1.71] | 1.46 [1.10-1.69] | 0.867 |
| **HF→δ** | 1.50 [1.08-1.71] | 1.49 [1.19-1.74] | 1.48 [0.93-1.72] | 0.436 |
| **HF→θ** | 1.47 [1.00-1.69] | 1.51 [1.07-1.77] | 1.41 [0.96-1.67] | 0.389 |
| **HF→α** | 1.41 [0.89-1.65] | 1.52 [1.14-1.80] | 1.38 [0.90-1.70] | 0.218 |
| ^1^Median [25%-75%] | | | | |
| ^2^Kruskal-Wallis rank sum test | | | | |

***Supplementary Table 2. Heart rate variability, EEG power and bilateral brain-heart interactions according to the standardized interpretation of EEG background following to the ACNS classification.***

|  | **SSEP N20** | |  |
| --- | --- | --- | --- |
| **Markers** | **N20 bilaterally absent, N = 32^1^** | **N20 uni-or bi-laterally present,  N = 80^1^** | **p-value^2^** |
| **ECG Heart Rate Variability** | | | |
| **LF (ms²/Hz)** | 0.04 [0-0.11] | 0.05 [0-0.22] | 0.175 |
| **HF (ms²/Hz)** | 0.02 [0.01-0.08] | 0.03 [0.01-0.16] | 0.320 |
| **LF/HF** | 0.71 [0.17-1.79] | 1.19 [0.32-2.68] | 0.148 |
| **EEG Power Spectral Density** | | | |
| **δ (V²/Hz)** | 4.7x10^-12^ [1.3x10^-12^-3.1x10^-11^] | 8.6x10^-11^ [1.5x10^-11^-2.5x10^-10^] | **1e-05** |
| **θ (V²/Hz)** | 1.9x10^-12^ [3.9x10^-13^-1.1x10^-11^] | 3.7x10^-11^ [6.5x10^-12^-1.1x10^-11^] | **2e-05** |
| **α (V²/Hz)** | 7.5x10^-13^ [1.4x10^-13^-3.7x10^-12^] | 1.0x10^-11^ [2.0x10^-12^-2.9x10^-11^] | **1e-04** |
| **Brain-to-heart coupling strength (median)** | | | |
| **δ→LF** | -1.3x10^10^ [-4.5x10^10^--8.9x10^8^] | -7.3x10^8^ [-4.3x10^9^--9.2x10^7^] | **1e-03** |
| **θ→LF** | -3.6x10^10^ [-1.6x10^11^--1.7x10^9^] | -1.9x10^9^ [-1.8x10^10^--2.0x10^8^] | **0.002** |
| **α→LF** | -5.3x10^10^ [-5.0x10^11^--3.5x10^9^] | -6.5x10^9^ [-5.3x10^10^--5.9x10^8^] | **0.004** |
| **δ→HF** | 2.8x10^10^ [5.3x10^9^-8.3x10^10^] | 2.3x10^9^ [2.9x10^8^-1.1x10^10^] | **3e-05** |
| **θ→HF** | 8.7x10^10^ [1.0x10^10^-2.9x10^11^] | 3.5x10^9^ [7.2x10^8^-3.9x10^10^] | **6e-05** |
| **α→HF** | 1.7x10^11^ [4.3x10^10^-8.8x10^11^] | 1.3x10^10^ [2.6x10^9^-1.2x10^11^] | **3e-04** |
| **Brain-to-heart coupling complexity (RCMSE)** | | | |
| **δ→LF** | 1.10 [0.70-1.45] | 1.19 [0.51-1.57] | 0.745 |
| **θ→LF** | 1.02 [0.58-1.43] | 1.18 [0.52-1.55] | 0.526 |
| **α→LF** | 1.15 [0.73-1.48] | 1.24 [0.51-1.60] | 0.577 |
| **δ→HF** | 0.77 [0.50-1.28] | 1.19 [0.35-1.54] | 0.600 |
| **θ→HF** | 0.76 [0.45-1.21] | 1.23 [0.45-1.55] | 0.222 |
| **α→HF** | 0.83 [0.48-1.34] | 1.22 [0.44-1.54] | 0.317 |
| **Heart-to-brain coupling strength (median)** | | | |
| **LF→δ** | 1.8x10^-11^ [5.7x10^-12^-1.5x10^-10^] | 2.7x10^-10^ [3.5x10^-11^-1.7x10^-9^] | **0.002** |
| **LF→θ** | 8.3x10^-12^ [1.1x10^-12^-8.7x10^-11^] | 1.2x10^-10^ [8.8x10^-12^-7.8x10^-10^] | **0.003** |
| **LF→α** | 3.1x10^-12^ [5.1x10^-13^-2.1x10^-11^] | 3.6x10^-11^ [3.2x10^-12^-2.3x10^-10^] | **0.007** |
| **HF→δ** | 1.1x10^-11^ [2.2x10^-12^-1.1x10^-10^] | 1.9x10^-10^ [1.7x10^-11^-1.0x10^-9^] | **0.002** |
| **HF→θ** | 3.4x10^-12^ [8.2x10^-13^-5.7x10^-11^] | 8.6x10^-11^ [5.5x10^-12^-6.7x10^-10^] | **0.004** |
| **HF→α** | 1.8x10^-12^ [5.3x10^-13^-1.8x10^-11^] | 2.2x10^-11^ [1.5x10^-12^-2.1x10^-10^] | **0.007** |
| **Heart-to-brain coupling complexity (RCMSE)** | | | |
| **LF→δ** | 1.52 [1.11-1.75] | 1.26 [1.12-1.56] | 0.097 |
| **LF→θ** | 1.49 [1.18-1.75] | 1.36 [1.04-1.57] | 0.148 |
| **LF→α** | 1.49 [1.24-1.73] | 1.42 [1.01-1.63] | 0.298 |
| **HF→δ** | 1.48 [1.26-1.72] | 1.46 [0.99-1.66] | 0.289 |
| **HF→θ** | 1.58 [1.09-1.72] | 1.43 [1.05-1.75] | 0.702 |
| **HF→α** | 1.42 [1.02-1.59] | 1.44 [1.01-1.76] | 0.489 |
| ^1^Median [25%-75%] | | | |
| ^2^Wilcoxon rank sum test | | | |

***Supplementary Table 3. Heart rate variability, EEG power and bilateral brain-heart interactions according to the N20 SSEP results.***

2. ***ECG heart-rate variability and EEG power spectral densities according to neurological outcome (Supplementary table 4)***

|  | **Best CPC at 3 months** | |  |
| --- | --- | --- | --- |
| **Characteristic** | **Good, N = 47^1^** | **Poor, N = 134^1^** | **p-value^2^** |
| **ECG Heart Rate Variability** | | | |
| **LF (ms²/Hz)** | 0.08 [0.02-0.41] | 0.05 [0.01-0.17] | 0.062 |
| **HF (ms²/Hz)** | 0.05 [0.01-0.16] | 0.03 [0.01-0.15] | 0.442 |
| **LF/HF** | 1.89 [0.76-3.99] | 0.97 [0.30-2.44] | **0.017** |
| **EEG Power Spectral Densities** | | | |
| **δ (V²/Hz)** | 6e-11 [2.9e-11-1.6e-10] | 6e-11 [4.0e-12-2.5e-10] | 0.137 |
| **θ (V²/Hz)** | 3.7e-11 [2.1e-11-6.1e-11] | 1.6e-11 [1.5e-12-1.2e-10] | 0.057 |
| **α (V²/Hz)** | 1.5e-11 [5.4e-12-2.9e-11] | 5.0e-12 [5.4e-13-2.9e-11] | **0.020** |

***Supplementary Table 4. ECG heart-rate variability and EEG power spectral densities according to patient’s neurological outcome.***

*Abbreviations: HF: high frequency, Hz: Hertz, LF: low frequency, ms: millisecond, p: p-value, V: volt.*

3. ***Brain-to-heart strength and complexity according to previous beta-blocker exposure and to sedation during EEG recording (Supplementary tables 5 and 6)***

|  | **Previous betablocker exposure** | |  |
| --- | --- | --- | --- |
| **Characteristic** | **No, N = 137^1^** | **Yes, N = 41^1^** | **p-value^2^** |
| **Brain-to-heart coupling strength (median)** | | | |
| **δ→LF** | -6.7x10^8^ [-5.5x10^9^--9.1x10^7^] | -1.5x10^9^ [-8.8x10^9^--1.3x10^8^] | 0.429 |
| **θ→LF** | -1.5x10^9^ [-2.1x10^10^--1.4x10^8^] | -2.8x10^9^ [-2.9x10^10^--2.4x10^8^] | 0.344 |
| **α→LF** | -4.1x10^9^ [-6.2x10^10^--3.9x10^8^] | -1.1x10^10^ [-1.0x10^11^--1.2x10^9^] | 0.275 |
| **δ→HF** | 1.9x10^9^ [4.5x10^8^-1.7x10^10^] | 2.7x10^9^ [4.8x10^8^-1.7x10^10^] | 0.774 |
| **θ→HF** | 4.4x10^9^ [7.7x10^8^-4.3x10^10^] | 7.4x10^9^ [5.9x10^8^-7.8x10^10^] | 0.571 |
| **α→HF** | 1.4x10^10^ [1.7x10^9^-1.4x10^11^] | 4.6x10^10^ [3.5x10^9^-2.0x10^11^] | 0.407 |
| **Brain-to-heart coupling complexity (RCMSE)** | | | |
| **δ→LF** | 1.14 [0.48-1.65] | 1.20 [0.85-1.60] | 0.302 |
| **θ→LF** | 1.10 [0.51-1.68] | 1.28 [0.81-1.55] | 0.276 |
| **α→LF** | 1.16 [0.51-1.64] | 1.28 [0.89-1.56] | 0.496 |
| **δ→HF** | 1.08 [0.37-1.52] | 1.10 [0.52-1.52] | 0.631 |
| **θ→HF** | 1.10 [0.44-1.54] | 1.19 [0.53-1.54] | 0.573 |
| **α→HF** | 1.07 [0.44-1.54] | 1.19 [0.51-1.54] | 0.753 |
| ^1^n (%); Median [25%-75%] | | | |
| ^2^Pearson's Chi-squared test; Wilcoxon rank sum test | | | |

***Supplementary Table 5. Brain-to-heart coupling strength and complexity according to betablocker exposure prior to ICU hospitalization.***

|  | **Sedation during EEG** | |  |
| --- | --- | --- | --- |
| **Characteristic** | **No, N = 131^1^** | **Yes, N = 48^1^** | **p-value^2^** |
| **Brain-to-heart coupling strength (median)** | | | |
| **δ→LF** | -7.6x10^8^ [-6.9x10^9^--1.2x10^8^] | -6.0x10^8^ [-4.3x109--3.3x10^7^] | 0.527 |
| **θ→LF** | -1.7x10^9^ [-2.3x10^10^--1.9x10^8^] | -1.6x10^9^ [-2.0x10^10^--4.2x10^7^] | 0.542 |
| **α→LF** | -4.7x10^9^ [-6.2x10^10^--5.1x10^8^] | -6.2x10^9^ [-9.2x10^10^--6.4x10^7^] | 0.407 |
| **δ→HF** | 2.4x10^9^ [4.0x10^8^-1.7x10^10^] | 1.8x10^9^ [6.3x10^8^-1.1x10^10^] | 0.753 |
| **θ→HF** | 4.7x10^9^ [6.6x10^8^-5.7x10^10^] | 5.8x10^9^ [7.4x10^8^-4.5x10^10^] | 0.970 |
| **α→HF** | 2.1x10^10^ [2.9x10^9^-1.8x10^11^] | 2.4x10^10^ [1.2x10^9^-1.6x10^11^] | 0.533 |
| **Brain-to-heart coupling complexity (RCMSE)** | | | |
| **δ→LF** | 1.19 [0.51-1.61] | 1.01 [0.55-1.58] | 0.565 |
| **θ→LF** | 1.20 [0.53-1.60] | 1.02 [0.58-1.56] | 0.527 |
| **α→LF** | 1.23 [0.51-1.64] | 1.03 [0.58-1.60] | 0.645 |
| **δ→HF** | 1.10 [0.46-1.52] | 0.88 [0.40-1.52] | 0.736 |
| **θ→HF** | 1.19 [0.49-1.53] | 0.94 [0.44-1.56] | 0.503 |
| **α→HF** | 1.11 [0.49-1.54] | 0.94 [0.45-1.51] | 0.680 |
| ^1^Median [25%-75%] | | | |
| ^2^Wilcoxon rank sum test | | | |

***Supplementary Table 6. Brain-to-heart coupling strength and complexity according to the infusion of sedation during EEG acquisition.***

4. ***Prognostic performances of ECG heart-rate variability, EEG power spectral densities and brain-heart interactions markers (Supplementary Table 7)***

|  | **Sen (%)** | **Spe (%-** | **PPV (%)** | **NPV (%)** | **AUC** |
| --- | --- | --- | --- | --- | --- |
| **ECG Heart Rate Variability** | | | | | |
| **LF (ms²/Hz)** | 71 [36-96] | 55 [21-85] | 81 [77-89] | 39 [30-70] | 0.59 [0.49-0.69] |
| **HF (ms²/Hz)** | 63 [7-79] | 57 [36-100] | 81 [76-100] | 33 [27-44] | 0.54 [0.44-0.63] |
| **LF/HF** | 68 [28-92] | 60 [28-91] | 83 [77-92] | 39 [30-59] | 0.62 [0.52-0.71] |
| **EEG Power Spectral Densities** | | | | | |
| **δ (V²/Hz)** | 31 [5-54] | 83 [60-100] | 84 [77-100] | 30 [27-34] | 0.43 [0.34-0.51] |
| **θ (V²/Hz)** | 46 [30-58] | 91 [79-100] | 94 [88-100] | 37 [33-42] | 0.59 [0.51-0.67] |
| **α (V²/Hz)** | 48 [28-66] | 85 [66-98] | 90 [84-98] | 36 [31-43] | 0.61 [0.53-0.70] |
| **Brain-to-heart coupling strength (median)** | | | | | |
| **δ→LF** | 60 [24-97] | 72 [28-100] | 86 [79-100] | 38 [31-76] | 0.67 [0.58-0.76] |
| **θ→LF** | 55 [25-90] | 79 [38-100] | 88 [80-100] | 38 [31-58] | 0.67 [0.59-0.76] |
| **α→LF** | 67 [25-90] | 66 [38-98] | 85 [80-97] | 41 [31-55] | 0.66 [0.58-0.75] |
| **δ→HF** | 41 [24-59] | 91 [72-100] | 92 [85-100] | 35 [31-40] | 0.60 [0.52-0.69] |
| **θ→HF** | 42 [25-65] | 89 [66-100] | 92 [84-100] | 35 [31-42] | 0.62 [0.53-0.70] |
| **α→HF** | 51 [21-75] | 79 [51-100] | 87 [80-100] | 36 [30-47] | 0.62 [0.53-0.70] |
| **Brain-to-heart coupling complexity (RCMSE)** | | | | | |
| **δ→LF** | 41 [24-90] | 85 [32-98] | 88 [78-97] | 34 [30-55] | 0.61 [0.52-0.71] |
| **θ→LF** | 43 [26-91] | 83 [32-96] | 87 [78-97] | 35 [30-61] | 0.62 [0.53-0.71] |
| **α→LF** | 54 [27-90] | 72 [30-96] | 84 [78-96] | 36 [30-58] | 0.60 [0.51-0.70] |
| **δ→HF** | 66 [23-86] | 64 [36-96] | 84 [78-96] | 38 [30-52] | 0.62 [0.53-0.71] |
| **θ→HF** | 54 [23-85] | 72 [36-98] | 85 [79-97] | 36 [30-50] | 0.61 [0.52-0.70] |
| **α→HF** | 63 [23-80] | 64 [40-96] | 83 [78-93] | 37 [29-48] | 0.60 [0.51-0.69] |
| **Heart-to-brain coupling strength (median)** | | | | | |
| **LF→δ** | 53 [6-92] | 64 [19-100] | 81 [76-100] | 31 [27-45] | 0.51 [0.42-0.61] |
| **LF→θ** | 32 [10-72] | 85 [43-100] | 85 [77-100] | 30 [27-39] | 0.53 [0.43-0.62] |
| **LF→α** | 34 [12-74] | 83 [40-98] | 84 [77-97] | 31 [27-38] | 0.53 [0.44-0.62] |
| **HF→δ** | 63 [9-78] | 57 [40-100] | 81 [77-100] | 35 [28-44] | 0.54 [0.45-0.64] |
| **HF→θ** | 57 [11-82] | 64 [34-100] | 82 [77-100] | 33 [28-44] | 0.54 [0.45-0.64] |
| **HF→α** | 47 [13-86] | 72 [30-100] | 83 [77-100] | 32 [28-43] | 0.54 [0.45-0.64] |
| **Heart-to-brain coupling complexity (RCMSE)** | | | | | |
| **LF→δ** | 56 [7-97] | 62 [11-100] | 81 [75-100] | 32 [27-60] | 0.53 [0.43-0.62] |
| **LF→θ** | 51 [18-87] | 70 [32-96] | 83 [77-94] | 34 [29-49] | 0.58 [0.49-0.67] |
| **LF→α** | 46 [14-80] | 72 [36-98] | 82 [76-97] | 32 [27-43] | 0.54 [0.45-0.63] |
| **HF→δ** | 56 [4-96] | 60 [13-100] | 80 [75-100] | 32 [27-50] | 0.50 [0.40-0.60] |
| **HF→θ** | 31 [9-91] | 85 [21-100] | 84 [76-100] | 30 [27-48] | 0.50 [0.41-0.60] |
| **HF→α** | 73 [7-94] | 47 [17-100] | 80 [76-100] | 35 [27-54] | 0.53 [0.44-0.63] |

***Supplementary Table 7. Prognostic performances of the different ECG, EEG and brain-heart markers for poor neurological outcome***

*Sensitivity (Sen), specificity (Spe), positive predictive value (PPV) and negative predictive value (NPV) were computed for the prediction of poor neurological outcome according to the best threshold of the area under the ROC curve (AUC) based on the Youden index. All values are expressed as % and their 95% confidence interval.*

5. ***Heartbeat Evoked Potentials according to outcome (Supplementary Figure 2)***

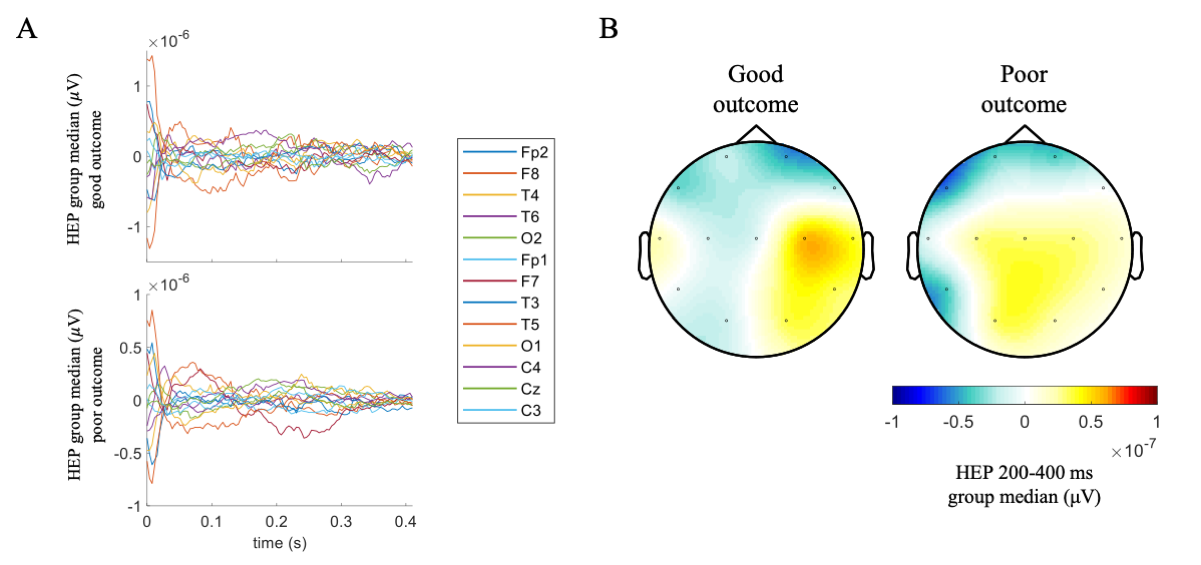

***Supplementary Figure 2. Heartbeat Evoked Potentials according to patients’ neurological outcome. (A)*** *HEP time course with respect to the R-peak.* ***(B)*** *HEP scalp topography distribution in the interval 200-400 ms with respect to the R-peak. Non-significant differences are found between the patients’ groups, as performed by cluster permutation analyses.*
